# Can we clinically identify pre-symptomatic and asymptomatic COVID-19?

**DOI:** 10.1101/2021.07.23.21260676

**Authors:** Salaheldin Elhamamsy, Frank DeVone, Tom Bayer, Chris Halladay, Marilyne Cadieux, Kevin McConeghy, Ashna Rajan, Moniyka Sachar, Nadia Mujahid, Aman Nanda, Lynn McNicoll, James L Rudolph, Stefan Gravenstein

## Abstract

**Objectives:** COVID-19 has had a severe impact on morbidity and mortality among nursing home (NH) residents. Earlier detection of SARS-CoV-2 may position us to better mitigate risk of spread. Both asymptomatic or pre-symptomatic transmission are common in outbreaks, and threshold temperatures, such as 38C, for screening for infection could miss timely detection in the majority.

**Design:** Retrospective cohort study using electronic health records

**Methods:** We hypothesized that in long-term care residents, temperature trends with SARS-CoV-2 infection could identify infection in pre-symptomatic and asymptomatic individuals earlier. We collected information about age and other demographics, baseline temperature, and specific comorbidities. We created standardized definitions, and an alternative hypothetical model to test measures of temperature variation and compare outcomes to the VA reality.

**Settings and participants:** Our subjects were 6,176 residents of the VA NHs who underwent SARS-CoV-2 trigger testing.

**Results:** We showed that a change from baseline of >0.4C identifies 47% of the SARS-CoV-2 positive NH residents early, and achieves earlier detection by 42.2 hours. Range improves early detection to 55% when paired with a 37.2C cutoff, and achieves earlier detection by 44.4 hours. Temperature elevation >0.4C from baseline, when combined with a 0.7C range, would detect 52% early, leading to earlier detection by more than 3 days in 22% of the residents. This earlier detection comes at the expense of triggering 57,793 tests, as compared to the number of trigger tests ordered in the VA system of 40,691.

**Conclusion and implications:** Our model suggests that current clinical screening for SARS-CoV-2 in NHs can be substantially improved upon by triggering testing using a patient-derived baseline temperature with a 0.4C degree relative elevation or temperature variability of 0.7C trigger threshold for SARS-CoV2 testing. Such triggers could be automated in facilities that track temperatures in their electronic records.

## Introduction

Early detection of SARS-CoV-2 allows interventions that can limit severity of disease and, in closed settings such as NHs, can mitigate risk of spread within the population. SARS-CoV-2 has a severe impact on morbidity and mortality among NH residents (4); accordingly, early detection has become a key strategy to reduce morbidity and mortality.

Current strategies for early detection include systematic temperature and symptom screening for SARS-CoV-2 infection. However, several investigations of SARS-CoV-2 outbreaks in NHs show that asymptomatic or pre-symptomatic transmission is common (3), and by the time clinical screening identifies infected individuals, infection has already spread to other residents and/or healthcare workers. This observation has prompted a second strategy, in which the entire population of healthcare workers and residents of a long-term care facility undergo testing, also called mass testing, or sweep testing. In US nursing homes (NHs), mass testing has been required in some states or individual facilities when community SARS-CoV-2 infection rates have been high, even as often as twice weekly.

This approach to disease prevention via late disease detection results in enforced isolation of residents and increased use of PPE, generating a human and financial toll. Systematic clinical screening is time consuming, and frequent mass testing is expensive. An important question this observation poses is what alternative strategies might allow us to do less frequent mass testing or only clinically triggered testing without sacrificing early detection.

Because daily temperature monitoring commonly has been used to screen NH residents for SARS-CoV-2 infection, we hypothesized that early temperature trends in the course of SARS-CoV-2 infection could identify either or both pre-symptomatic and asymptomatic individuals in the long-term care setting.

The Veterans Health Administration (VA) mandated daily clinical and temperature screening of all residents of its Community Living Centers, beginning March 1, 2020, like non-VA NHs. Additionally, along with systematic testing based on clinical screening, the VA required all its NH residents to be mass tested starting April 10th. These clinical and laboratory data are captured in the VA’s electronic health records, allowing evaluation of temperature trends in individuals who did and did not have SARS-CoV-2 infection.

## Methods

The study was conducted in the Veterans Affairs (VA) 133 NHs in the United States. The VA oversees care in these facilities using a centralized computerized patient record system (CPRS) that stores all clinical and laboratory data, making them available for analysis. Included are the data for SARS-CoV-2 daily screening and testing results. This study was approved by the Providence Veterans Administration Medical Center’s Institutional Review Board.

Our subjects included all residents of the VA NHs who were living there at any time between March 1, 2020, to December 13, 2020, and underwent SARS-CoV-2 testing during their stay. We excluded subjects whose most recent test we estimated to be a non-clinically triggered test (Mass test).

Body temperature was determined by several methods during routine nursing care; each CLC uses standard equipment to measure temperature and enters the reading into the electronic medical record.

We created an alternative hypothetical scenario to model what would have happened if different metrics were used to screen residents for SARS-CoV-2 and compared it to what really happened in the VA (VA reality). We defined three metrics by which a resident could trigger a SARS-CoV-2 test. 1) A simple T-max cutoff, if a resident’s one time temperature measurement exceeded a value, then it would trigger a SARS-CoV-2 test. 2) An elevation from baseline cutoff; If a resident’s temperature exceeded their base temperature by a declared amount, then it would trigger a SARS-CoV-2 test. 3) A Temperature Range cutoff or temperature variability; within a three-day period, if the difference between the maximum temperature on and the minimum temperature exceeds a declared threshold value, then a SARS-CoV-2 test would be triggered. SARS-CoV-2 date was defined as the day of a resident’s positive test or, if they never have had a positive test in our study period, the sampling date of their last negative SARS-CoV-2 test. SARS-CoV-2 Period was defined as 14 days before and after their SARS-CoV-2 date. Baseline temperature was defined as the mean of the first 5 temperatures on record before a resident’s SARS-CoV-2 period. We used the first daily temperature occurring after 4 am.

We calculated baseline temperatures by averaging the first 5 daily temperatures recorded before the SARS-CoV-2 date. We identified SARS-CoV-2 polymerase chain reaction (PCR’) testing results from the VA’s electronic medical records. The VA developed a harmonized definition of SARS-CoV-2 test results, requiring a PCR test from a certified laboratory.

Mass testing or Sweep Test is testing for routine infection control purposes, or mass testing of large amounts of residents due to COVID-19 protocols as determined by the VA. Clinically Triggered tests (Trigger test) are defined as tests based on clinical signs or symptoms. We developed an algorithm to determine testing routine based on density of testing on a given day per facility. If a facility had a density of testing larger than its study periods mean + 1-SD, or if a facility exceeded 30% residents having a test in one day, that would be considered mass testing.

We also defined 4 measures to determine the performance of our possible SARS-CoV-2 triggers. Earlier detection per resident was measured in hours; the mean time a resident was detected earlier using this metric as compared to VA reality. Among our subjects, the actual percentage of positive cases detected early using this metric was defined as the infected identified early. For all residents, the total time of earlier detection using a particular metric as compared to VA reality was defined as cumulative days earlier detection. The total number of tests triggered by our subjects using these metrics was defined as tests triggered.

The total number of SARS-CoV-2 tests (clinically triggered and mass testing) among all residents in the VA NHs during the study period was 123,288. We compared that to the total number of tests triggered by our cohort using our metrics. To compare, we reduced our sample to residents with temperature data on record and who was last tested using a clinically triggered test. In addition, if found positive, we excluded future tests on record. We estimated the number of SARS-CoV-2 clinically triggered tests among this cohort to be 40,691. We estimate the number of SARS-CoV-2 mass tests to be 82,597. Once a resident triggers a SARS-CoV-2 test and is negative, they cannot trigger a SARS-CoV-2 for 7 days. In addition, for this analysis, we assume once positive, a resident cannot again be positive within the study period. If a resident was not detected within 7 days of when positive, we assume that they would have been found through other methods. We also allow for combinations of these three metrics, in which any of them could trigger a SARS-CoV-2 test, or we require multiple metrics to be met for a SARS-CoV-2 test to trigger. Additionally, any resident who has a temperature >38 °C will also trigger a test. We used these metrics to see if we could determine SARS-CoV-2 positivity prior to VA reality ‘‘early detection’’ and look at the ‘‘cost’’ number of tests triggered.

We used standard deviation, p-value (with an alpha set at 0.05) and confidence intervals to compare groups. Statistical analyses included R.4.0.2 and Microsoft SQL Server 2017. For the demographics table, continuous variables were confirmed with an ANOVA test; categorical variables were compared with χ2.

## Results

We collected information about age and other demographics, baseline temperature, and specific comorbidities (Table 1). The 133 CLCs admitted 15,043 residents during the study period. For our study, we evaluated the 6176 subset whose tests for SARS-CoV-2 were ordered by clinical indication, i.e., trigger tested. Those in whom SARS-CoV-2 testing confirmed infection were older (74.27 vs. 71.40 years, *P* < 0.0014) than those with tests that did not confirm infection.

**Table 1.** Demographics.

|  | <u>Total</u> | <u>COV+</u> | <u>COV-</u> | <u>P-Value</u> |
| --- | --- | --- | --- | --- |
| N | 6,176 | 914 | 5,262 | N/A |
| Age | 71.82 (11.6) | 74.27 (10.87) | 71.40 (11.66) | < 0.001 |
| Male | 5,913 (95.74%) | 888 (97.16%) | 5,025 (95.50%) | 0.028 |
| Race: White | 4,391 (71.10%) | 616 (67.40%) | 3,775 (71.74%) | 0.008 |
| Race: Black | 1,330 (21.53%) | 225 (24.62%) | 1,105 (21.00%) | 0.016 |
| Race: Other | 455 (7.37%) | 73 (7.99%) | 382 (7.26%) | 0.479 |
| Diabetes Mellitus (DM) | 2,621 (42.45%) | 395 (43.22%) | 2,226 (42.31%) | 0.6349 |
| DMcx | 2,779 (45.00%) | 398 (43.54%) | 2,381 (45.26%) | 0.3551 |
| Hypertension (HT) | 4,719 (76.42%) | 693 (75.82%) | 4,026 (76.53%) | 0.6737 |
| HTNcx | 2,673 (43.29%) | 364 (39.82%) | 2,309 (43.89%) | 0.0243 |
| Congestive Heart Failure | 2,138 (34.62%) | 286 (31.29%) | 1,852 (35.20%) | 0.0240 |
| Pulmonary | 2,567 (41.57%) | 363 (39.72%) | 2,204 (41.89%) | 0.2314 |
| BMI | 28.27 (7.37) | 28.24 (7.33) | 28.28 (7.37) | 0.4513 |
| Valvular | 845 (13.68%) | 93 (10.18%) | 752 (14.29%) | 0.0010 |
| Alcohol | 1,107 (17.93%) | 146 (15.97%) | 961 (18.27%) | 0.1005 |
| Drugs | 882 (14.28%) | 114 (12.47%) | 768 (14.60%) | 0.1002 |
| Anemia | 3,270 (52.96%) | 430 (47.05%) | 2,840 (53.98%) | 0.001 |
| Depression | 3,007 (48.70%) | 426 (46.61%) | 2,581 (49.06%) | 0.183 |
| Tumor | 1,244 (20.15%) | 152 (16.63%) | 1,092 (20.76%) | 0.0047 |
| Psychoses | 1,613 (26.12%) | 339 (37.09%) | 1,274 (24.22%) | <0.001 |
| TBI | 403 (6.53%) | 62 (6.78%) | 341 (6.48%) | 0.7884 |
| Baseline Temperature | 36.59 (0.24) | 36.58 (0.22) | 36.59 (0.24) | 0.9903 |

Temperatures with SARS-CoV-2 confirmed infections began rising as early as 7 days before testing and remained elevated as late as a 14-day follow-up. Among those residents with SARS-CoV-2, only 47.7% eventually met the fever threshold of 38°C. In our hypothetical model we used only temperature as a screening tool to determine whether, on average, we can identify those with infection earlier and compared this to the standard of care approach that really happened in the VA NHs; i.e., the “VA Reality.” Table 2 contains a highlight of possible positive SARS-CoV-2 detection methods (this is also displayed graphically in figure 1).

**Table 2:**
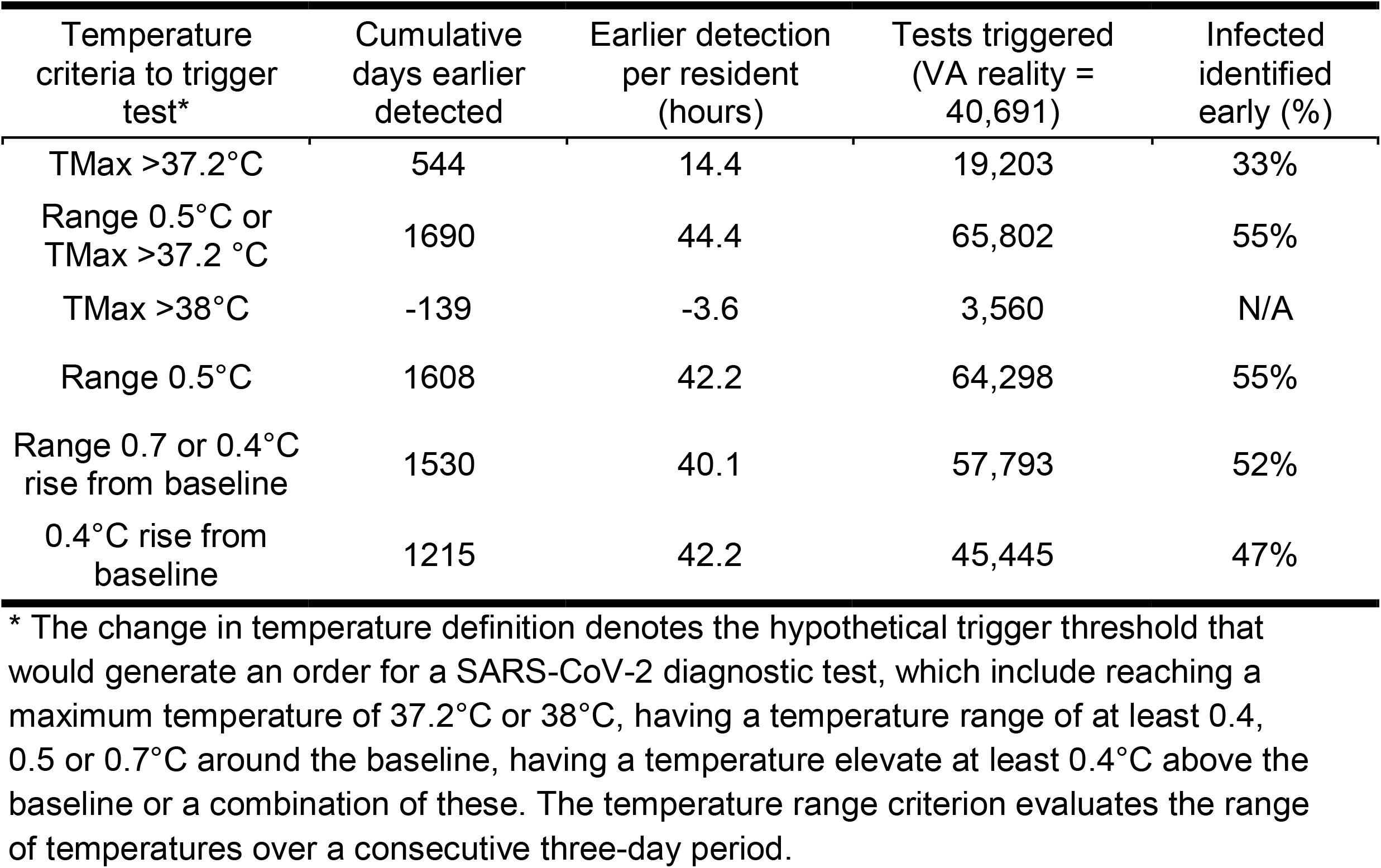
Relationship of temperature and time to SARS-CoV-2 detection compared to the “VA Reality.”

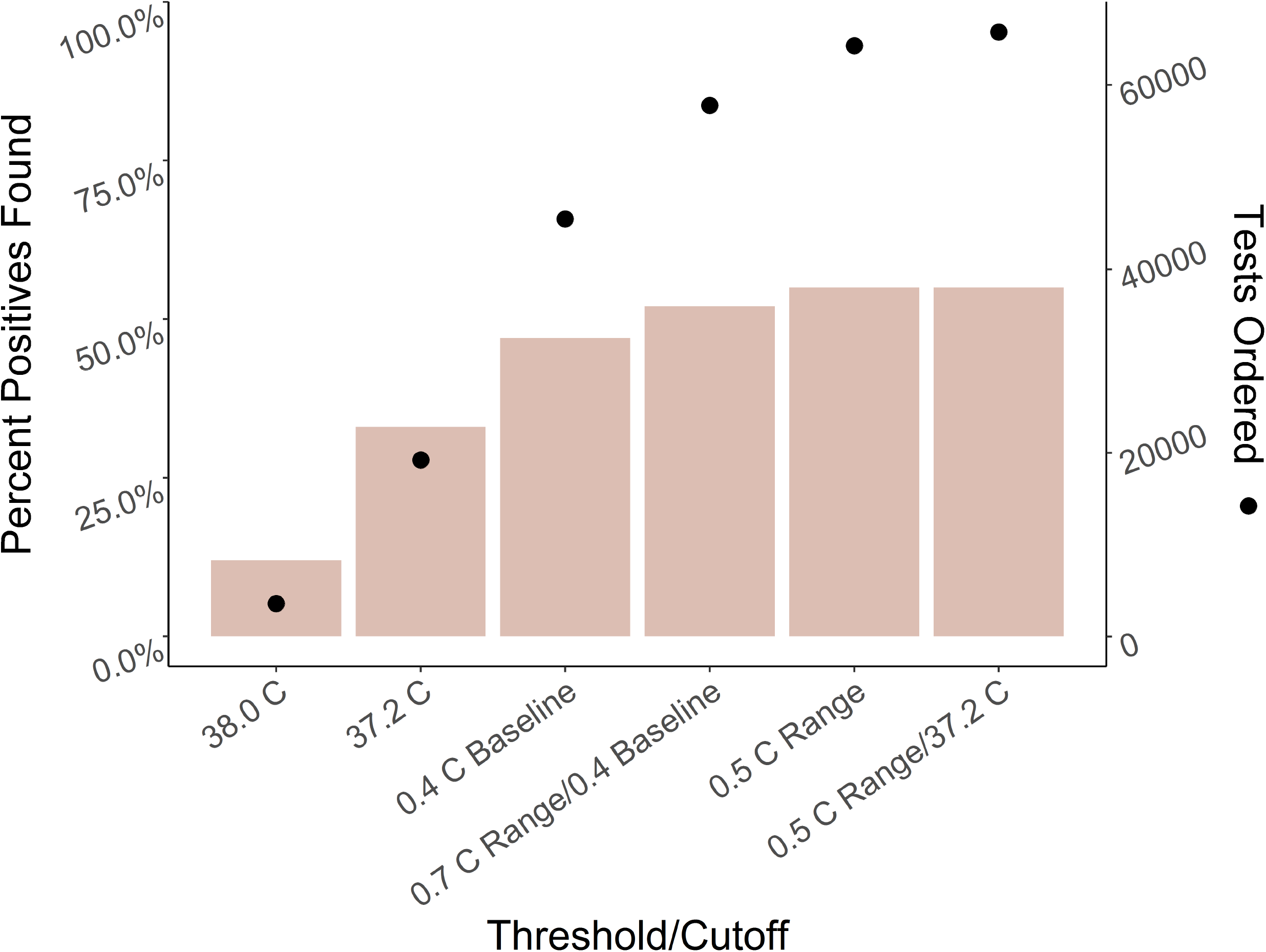

When we use a temperature threshold of 38.0°C to trigger a screening test for SARS-CoV-2 in NH residents, we will identify 39% of the cases. A temperature threshold of 38.0°C would trigger 3,575 tests but detect individual cases on average 3.6 hours/resident later compared to the VA Reality. When we use the lower temperature threshold of 37.2°C, we would detect 33% of the SARS-CoV-2 infected residents early, and on average detect those with infection 14.4 hours earlier.

Temperature elevation >0.4°C from baseline identifies 47% of residents with SARS-CoV-2 early. This threshold results in earlier detection by an average of about 42 hours per resident. Temperature variability increases with infection; using a range to select who to test improves early detection to 55% of those in whom SARS-CoV-2 was confirmed when paired with a simple 37.2°C cutoff, this improved early detection by about another 2.4 hours on average (f 44.4) hours per resident. This temperature cutoff would trigger 65,802 tests. With a variability of 0.5°C, we identify 55% of those infected early and achieve earlier identification of whom to test by an average of 42 hours per resident. This triggers 64,298 tests, some 24,000 more than the 40,691 tests ordered in the VA Reality.

Temperature elevation >0.4°C from baseline combined with a 0.7°C range would detect 52% of the SARS-CoV-2 infected residents early leading to earlier detection of 40 hours on average and would trigger 57,793 tests as compared to the number of trigger tests ordered in the VA system of 40,691. When we look at residents who are detected 24 hours or more prior to VA reality using this method we find that 50% of those infected with SARS-CoV-2 are detected one calendar day or more earlier than the VA reality (see figure 2), >25% are detected 4 or more days earlier, and 11.5% are detected 6 or more days earlier than VA reality.

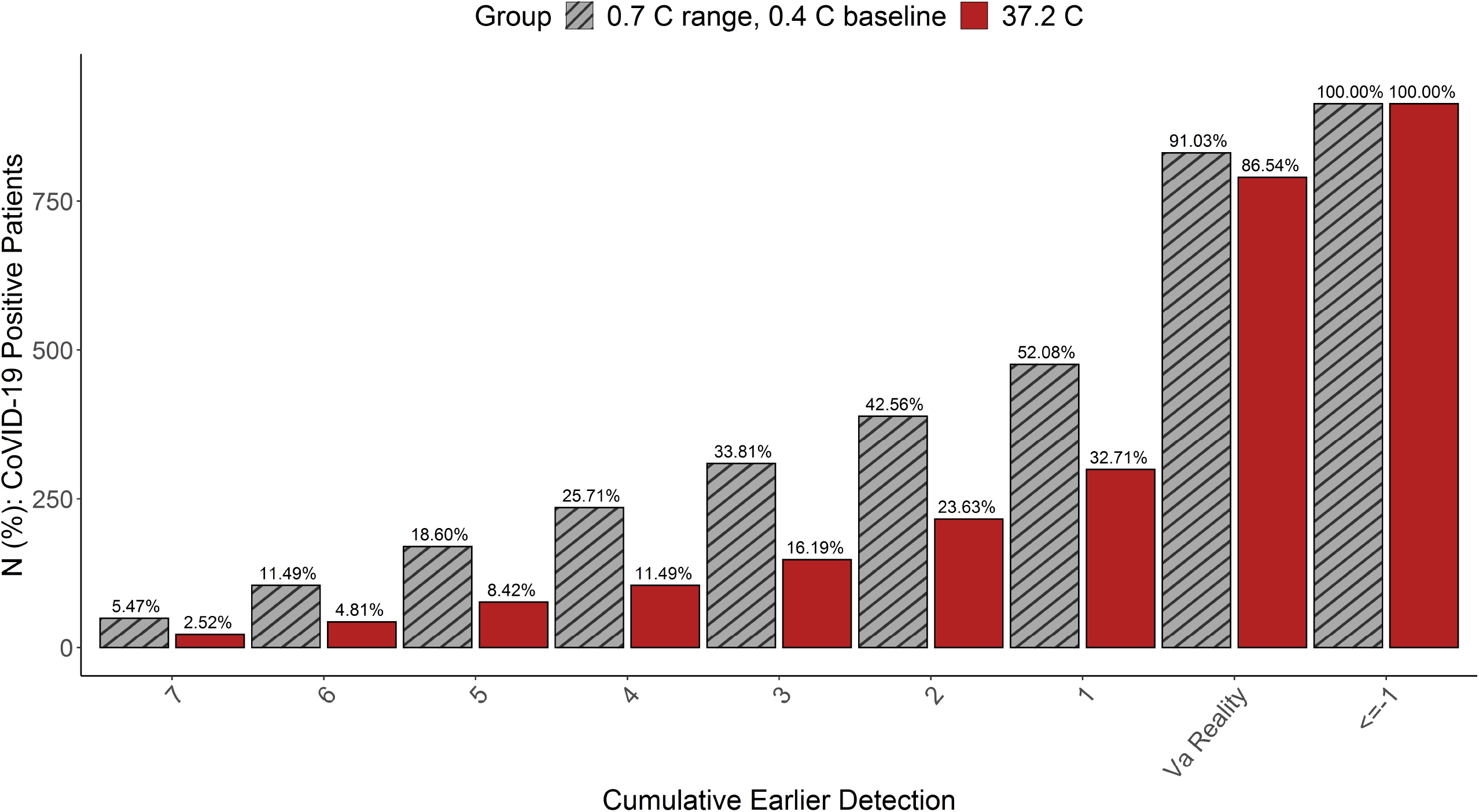

We performed a verification among the residents assumed to be sweep tested and found the simpler measures performed worse in every category, while the range scenarios performed better at the cost of a large increase in tests. If applying the range scenarios to large asymptomatic populations, caution, or the willingness to test at a large number must be considered.

## Discussion

Our data show that earlier detection of SARS-CoV-2 infection in the NH can be achieved using only individual or combined measures of temperature variability. Our hypothetical model using a 37.2°C temperature cutoff threshold compared to current practice demonstrates that the VA could identify individuals with SARS-CoV-2 for screening on average by 14.4 hours. This model could identify 32.71% of the cases one calendar day or more earlier, 16.2% are detected 3 days or more earlier and 11.4% are detected 4 days or more earlier than VA reality, which uses both clinical and temperature measures. The cost would be ordering 19,203 tests, versus 3,560 tests ordered when a temperature threshold of 38°C is used. It is important to know that in reality the VA ordered 40,691 clinically triggered tests for this cohort; temperature was not the only measure used to screen residents. Changing the temperature threshold to 37.2°C could give nursing facilities a major advantage in early detection of SARS-CoV-2; it would be easy to change current practice with little training.

The model that produces a superior predictive measure uses temperature variability and threshold - a 0.7°C range paired with a 0.4°C increase from baseline temperature. This model predicts a 40-hour average advantage in identifying those with SARS-CoV-2 infection for screening with 52% early detection, while triggering 57,793 tests. Not only does this approach identify over 50% of those infected with SARS-CoV-2 one calendar day or more earlier than the VA reality (see figure 2), but also over 25.7% are detected 4 or more days earlier and 11.5% are detected 6 or more days earlier than VA reality. This approach to detection could position facilities for substantially earlier outbreak control measures implementation and stopping the spread of SARS-CoV-2.

Individual temperatures provide incomplete clinical information and context. Our method leverages information already in the medical record to improve its utility for clinical decision making; if automated on the EHR platform, it could flag these outliers to reduce the burden tracking and calculating for busy medical providers. To help identify high risk individuals who would benefit from testing for SARS-CoV-2.

Older residents tend to have lower baseline temperatures (5), limiting utility of a simple temperature cutoff. Also, investigations of body temperature in older adults and NH residents have shown absolute temperature thresholds such as 38°C is insensitive. (4) NH residents can have atypical physiological responses to infection and might not be able to report symptoms if they have dementia.

SARS-CoV-2 infection can spread from residents who have not yet (presymptomatic) or will ever (asymptomatic) have symptoms, thus limiting effectiveness of clinical screening. As in our sample, clinical screening often has not identified or led to containment of SARS-CoV-2 spread in NHs early in the pandemic. The consequent expensive mass testing strategy has included weekly or sometimes bi-weekly testing of all residents and staff in a facility. The expense includes not only the cost of the test, but those attributable to staff time, resources, PPE consumption to conduct tests, and psychological cost for residents, families, and staff.

Using a person-derived baseline temperature by averaging the first daily temperatures over a 3–5-day period can solve this problem and is a more person-centered approach to defining fever. This concept can be used for other infectious diseases once it is validated.

Monitoring resident temperature is an inexpensive intervention that NH staff are well trained on. Understanding early temperature trends with SARS-CoV-2 infection can allow for earlier detection, better infection control, and cost reduction by transitioning away from mass testing strategies.

We noticed that using a threshold increase from baseline occurring in multiple readings offers a favorable balance of sensitivity and specificity relative to a single reading, but is a more complex method to implement in a clinical setting and will probably be done using the computer system.

As noted before, in total the VA ordered in the 133 NHs between clinically triggered tests and mass tests a total of 123,288 tests. Mass testing is an expensive non-directed way of testing residents in the NH for SARS-CoV-2 infection. As more data is available, we can transition to more directed testing.

On the facility scale, detection of clusters of residents in a nursing facility that trigger allows tracking and early intervention, a more efficient approach to infection control. Creating a centralized infection monitoring and control dashboard for the nursing facility makes sense. We see the potential for expanding this concept as we start collecting data with continuous temperature monitoring devices, a technology that already exists and is commercially available.

There are challenges to clinical implementation; to calculate the temperature range, several readings are needed, at a minimum a temperature per day over three days. Change from baseline requires an on-file measure of the resident’s baseline temperature. Finally, the temperature cutoff requires only a simple point measurement. Of the three proposed methods of early detection and their possible variants, range appears to offer the best early directions, but only when paired with the other measures. Performing these statistical analyses at the scale provided to us by the VA data allows us to better study the earlier detection of SARS-CoV-2 in a NH setting than we believe has previously been done. The strength of our analysis includes a large NH sample, frequent temperature monitoring during the pandemic in the VA system, and constant monitoring of COVID-19. The study population includes immune senescent older adults living in long term care facilities who are not often included in medical research.

Study limitations; our scenario is a hypothetical model, we only have point estimates for the temperatures, ideally we’d have continuous temperature monitoring to better understand the effect of SARS-CoV-2 on temperature over time. Our cohort is also older and mainly male and white (see table 1), we know these demographic variations can affect temperature. Our creation of a hypothetical model prevents us from performing more traditional statistical tests on our results, we created the measures discussed as our best substitute for more traditional sensitivity and specificity analysis. Some of these limitations could be overcome by using continuous temperature monitoring devices. Also, NHs typically do not have a documented baseline temperature defined for their residents. To do so, the EMR will need to be programmed to use existing temperature data that establish and track person-level baseline temperatures; then, it can also be set to alert the clinician to relevant changes from baseline.

Resident records did not distinguish whether clinical symptoms triggered the SARS-CoV-2 test. We therefore relied on a conservative definition for “trigger test” that limited our sample size by more than half. Had this information been available, we likely would have had a much larger portion of our sample available for the analysis.

A prospective study to test the prediction of the model is needed.

## Conclusions and implications

Our model suggests that current clinical screening for SARS-CoV-2 in NHs can be substantially improved upon by triggering testing using a resident-derived baseline temperature with a 0.4°C degree relative elevation or temperature variability of 0.7°C trigger threshold for SARS-CoV2 testing. This approach can lead to better infection control and reduce the need for mass testing in NHs. Such triggers could be automated in facilities that track temperatures in their electronic records. These data can be used for creating early detection algorithms that will be significantly enhanced when continuous temperature monitoring is available to high-risk NH residents.

## Data Availability

Data will be made available upon request

## Acknowledgement to contributors

We thank Dr Richard Besdine, MD, Alpert Medical School of Brown University, for final manuscript reviewing and editing.

